# Investigating group-based classes (‘weaning workshops’) to support complementary infant feeding in Irish primary care settings: A cross-sectional survey

**DOI:** 10.1101/2021.12.01.21267143

**Authors:** Caragh Flannery, Caroline Shea, Yvonne O’Brien, Joanne O’Halloran, Karen Matvienko-Sikar, Colette Kelly, Elaine Toomey

## Abstract

**Objective:** This study aims to 1) investigate current practice regarding ‘weaning workshops’ to support complementary infant feeding delivered within Irish primary care, 2) explore the experiences and opinions of community dietitians regarding optimal content and modes of delivery of weaning workshops, and 3) identify the key factors to be considered in the development and implementation of weaning workshops delivered within primary care.

**Design:** Cross-sectional survey

**Setting:** Irish primary care

**Participants:** 47 community-based dietitians

**Results:** Sixteen dietitians reported that workshops were run in their area with variable frequency, with ten reporting that workshops were never run in their area. Participants reported that mostly mothers of medium socioeconomic status attended weaning workshops when infants were aged between 4-7 months, and that feedback from workshop attendees was predominantly positive. Dietitians identified that key factors to be considered in future development and delivery of weaning workshops are 1) workshop characteristics such as content, timing and venue, 2) organisational characteristics such as availability of resources and multidisciplinary involvement, and 3) attendee characteristics such as socioeconomic status.

**Conclusions:** This study highlights substantial variability regarding provision of weaning workshops in Ireland, and a lack of standardisation regarding the provider, content, and frequency of workshops where workshops are being delivered. The study also provides unique insights into the experiences and opinions of primary care community dietitians regarding the development and delivery of weaning workshops in terms of optimal content and delivery options. These perspectives will make a valuable contribution given the dearth of evidence in this area internationally.

## Background

Optimum infant nutrition is a vital aspect of healthy child development and chronic disease prevention, and the complementary introduction of solid foods within the first two years of life plays a key role in this. International World Health Organisation (WHO) infant feeding guidelines recommend that infants be exclusively breastfed until 6 months, with nutritionally adequate and appropriate solid foods introduced from 6 months onwards (1, 2). However, despite these recommendations, a substantial proportion of infants globally are introduced to solids before four months (3, 4). In Ireland, a recent study identified that 18% of Irish infants had begun complementary feeding before 17 weeks, with another showing 13.5% taking solids between 12 and 16 weeks (5-7). Previous research also found that a high proportion of Irish infants aged six months consume foods high in energy, salt, saturated fats, and refined sugars (8, 9). Given the importance of establishing healthy dietary behaviours from childhood, supporting parents and primary caregivers to introduce solid foods for infants and young children in line with best practice recommendations is of key significance both nationally and internationally.

Multiple factors influence complementary infant feeding behaviours. For example, Fall et al (2011) identified several factors associated with the early introduction of solids, including younger maternal age, lower socioeconomic status (SES), lower maternal education, absent/short duration of breastfeeding and maternal smoking (10). More recently, Gutierrez-Camacho et al. (2019) identified that higher maternal education, higher age and higher household income are positively associated with healthy infant feeding behaviours (11). In addition, recent qualitative evidence syntheses explored parents’ perceptions and experiences of infant feeding and identified many factors influencing parents’ complementary feeding behaviours (12, 13). Both reviews found that complementary feeding behaviours are influenced by multiple sources of advice, in particular those of peers (i.e., family and friends) and healthcare professionals. The studies identified a need for greater education and support for parents regarding complementary feeding, particularly within primary care settings (13), and considering practical issues such as time and cost (12). The reviews also highlighted greater barriers within lower SES populations, such as the lack of affordability of healthy food (13), greater misconceptions and lack of knowledge (12).

Despite an increasing focus on the importance of appropriate complementary infant feeding, and the need to support parents and primary caregivers to engage in recommended behaviours, the existing evidence regarding the effectiveness of interventions to improve complementary feeding behaviours is inconsistent (12, 14, 15). Recent systematic reviews of infant feeding interventions have shown minimal effects on infant feeding practices and other important outcomes such as child weight (12, 14, 15). Methodological limitations of existing interventions have also been highlighted with poor application and/or use of behaviour change theory, a lack of systematic intervention development and a lack of focus on long-term implementation and sustainability (15-17). Results of these reviews have emphasised the importance of trustworthy, non-biased, consistent infant feeding information for parents while also facilitating the development of practical skills and providing hands-on support such as demonstrating how to prepare nutritional meals (12, 14-17). Given the important role that healthcare professionals can play in influencing infant feeding behaviours coupled with the strong social influence of family and friends (3, 12, 13), the current evidence therefore suggests a need for rigorously developed and evaluated sustainable community-based interventions that can be embedded into routine healthcare services. Primary care represents a promising setting for such interventions given its community-based context and an increasing global focus on health promotion and disease prevention within primary care practice (18-20). Given their expertise, community dietitians may have an important role to play in interventions that aim to improve healthy infant feeding; however, recent research has highlighted limited capacity amongst dietitians to prioritise preventive-focused interventions over complex clinical issues such as feeding challenges or problems (21).

Group-based parenting programmes have been previously recognised as an effective means to change parents’ behaviour, and when implemented in community-based settings, have been found to improve child social, emotional outcomes, as well as parenting skills (22-24). Furthermore, these group-based programmes have shown that they can be delivered with high levels of implementation fidelity and parental engagement (22, 25, 26). Other studies have identified factors which are associated with the success of parenting programmes including parental attitudes towards programme content, changes in parenting skills and confidence, and positive experiences of the group process (26-28). In the context of early child nutrition, group-based programmes focusing on complementary feeding, or ‘weaning workshops’, have been suggested as a promising approach to target infant feeding practices (12, 21, 23, 24, 29, 30). Such programmes are workshop-style events that can target several important aspects for effective community-based interventions, e.g. provide much needed access to evidence-based information, social support, and practical skills. However, apart from some notable exceptions (29, 30) there is limited research on the development, implementation and evaluation of weaning workshops within community or primary care settings, with the majority of evidence focusing predominantly on breastfeeding support or individually-delivered interventions (31, 32). Additionally, current practice surrounding weaning workshops and service offerings within routine care, and the optimum content and mode(s) of delivery have been poorly explored to date, both in Ireland and internationally. A previous unpublished report from the Irish Health Service Executive (HSE) in 2016 completed by 11 dietitians from 9 Irish community health organisation (CHO) areas identified that weaning workshops were being offered in 7/9 CHO areas, and that programme content and implementation was highly variable across settings (33). Ten of the included dietitians highlighted the need for standardised weaning workshop; however this report did not collect information from participants regarding their views on the optimum content or mode(s) of delivery of weaning workshops, nor the factors that need to be considered in their development and implementation in primary care settings. Due to the potential for community-based weaning workshops to support complementary infant feeding behaviours and the scant existing evidence internationally to inform appropriate development and implementation, further exploration of such interventions is needed.

This study aims to:

- Investigate current practice regarding the delivery of weaning workshops within Irish primary care settings,
- To explore the experiences and opinions of community dietitians regarding the optimal content and modes of delivery of weaning workshops, and
- To identify the key factors that should be considered to facilitate the development and implementation of effective and sustainable weaning workshops within routine primary care from the perspectives of community dietitians.

## Methods

### Study design

The research adopted a quantitative approach using a cross-sectional survey distributed to community-based dietitians working in the Irish primary healthcare service. We obtained ethical approval from the School of Public Health Internal Social Research Ethics Committee in University College Cork. The STROBE checklist for cross-sectional studies (34) was used to inform reporting of the findings (Appendix 1).

### Sampling and recruitment

All community-based HSE dietitians based in Ireland were eligible to participate in the study. After consultation with a senior community dietitian in primary care (JOH), it was established that there are approximately 90 community-based dietitians in Ireland were eligible to participate in the study. The survey was set up using ‘Lime Survey’, an online resource for hosting surveys and questionnaires. An email inviting all community-based HSE dietitians in Ireland was sent by a primary care dietitian manager (YOB) in the Cork area from May-June 2019. One reminder email was sent within this timeframe. The invitation email included an information sheet outlining the nature of the research, a link to the survey and instructions for obtaining consent. Before emailing, the survey was piloted with two participants (a researcher with a nutritional background and a dietitian), after which minor refinements were made to the wording of questions. The pilot responses were not included in the analysis.

### Data Collection

A 34-item survey was developed based on existing literature in conjunction with an unpublished report by the Health Service Executive (12, 29, 33, 35). In the first section, participants were asked to provide their age category, gender, CHO area, length of time qualified, whether they worked in primary care or health promotion and current position. The second section asked participants about current weaning workshops service offerings in their CHO area. Specifically, participants were asked questions about workshop characteristics including frequency, provider, content and resources required, and about attendee characteristics, e.g. numbers in attendance, average parent/caregiver type (e.g. mothers, fathers, grandparents etc), average socioeconomic status, average age of infant and their views on the general reception by attendees regarding the workshops. The next section asked participants about their specific experience and opinions of delivering weaning workshops, and their views of the factors influencing the development and delivery of weaning workshops and attendance by parents/caregivers. All of the above items were closed-ended questions with multiple-choice answers provided (n=30 items). Four open-ended questions were also included to capture participants’ views on 1) reasons that workshops are currently not available in their area (if applicable), 2) the most important factors to consider in development and delivery of weaning workshops, 3) recommendations regarding existing services and 3) any additional comments. We defined weaning workshops for participants as ‘any community-based, group-based classes that aim to address complementary feeding and the introduction of solid foods for parents and primary caregivers of infants and children’. The full questionnaire is provided in Appendix 2.

### Data analysis

The data were cleaned and then transferred into STATA (version 13) for analysis. A descriptive univariate analysis was conducted by CF for all dietitians’ characteristics, workshop characteristics and reported attendee characteristics, providing sample numbers and percentages. We used chi-squared test for categorical variables to present differences between responses from dietitians who had experience of delivering workshops and those who did not, but we did not calculate significance or p-values due to the small samples involved. All surveys were returned complete but scattered missing data reduced the sample size for some variables (further data provided on missing data in Appendix 3). In items that allowed for multiple responses (‘tick all that apply’), categories were created for most of the matching responses. For all additional responses, an ‘other’ category was created. Data from all four open-ended questions were analysed by ET using a conventional content analysis approach outlined by Hsieh and Shannon (2005) (36). This approach was chosen due to the likelihood of minimal depth and richness of responses. After reading and data familiarisation, ET highlighted text that captured key concepts and meaning from responses. These key concepts were then summarised, producing initial codes, which were refined and sorted into overarching categories which were subsequently discussed with CF.

## Results

### Dietitians’ characteristics

A total of 90 dietitians were invited to participate in this study; 47 (52%) consented to participate. Most participants were in the 31-40 years of age bracket (n=17, 39.5%). Community dietitians were predominantly female (n=41, 97.6%), based in primary care (n=33, 80.5%) and working as a dietitian for 1-5 years (n=14, 35%). Representation varied across the 9 CHO areas represented, ranging from 0 (CHO area 2) to 12 (28.6%) from CHO area 4. Most participants were in a senior dietitian role (n=31, 73.8%) (See Table 1).

**Table 1.** Dietitians’ characteristics.

| Variable | N (%) |
| --- | --- |
| **Age category (n=43)** |  |
| 18-30 years | 7 (16.3) |
| 31-40 years | 17 (39.5) |
| 41-50 years | 14 (32.6) |
| 51-60 years | 4 (9.3) |
| >60 years | 1 (2.3) |
| **Gender (n=42)** |  |
| Female | 41 (97.6) |
| Male | 1 (2.4) |
| **CHO (n=42)** |  |
| Area 1 | 3 (7.1) |
| Area 2 | 0 (0.0) |
| Area 3 | 3 (7.1) |
| Area 4 | 12 (28.6) |
| Area 5 | 4 (9.5) |
| Area 6 | 7 (16.7) |
| Area 7 | 5 (11.9) |
| Area 8 | 1 (2.4) |
| Area 9 | 7 (16.7) |
| **Length in profession (n=40)** |  |
| 1-5 years | 14 (35.0) |
| 6-10 years | 5 (12.5) |
| 11-15 years | 10 (25.0) |
| 16-20 years | 10 (25.0) |
| 21-25 years | 1 (2.5) |
| **Discipline (n=41)** |  |
| Primary care | 33 (80.5) |
| Health promotion | 4 (9.8) |
| Other | 4 (9.8) |
| **Current position (n=42)** |  |
| Dietitian manager | 3 (7.1) |
| Senior dietitian | 31 (73.81) |
| Staff-grade dietitian | 8 (19.1) |

CHO, Community Health Organisation; Area 1, Donegal, Sligo/Leitrim/West Cavan, Cavan/Monaghan LHOs; Area 2, Community Healthcare West; Area 3, Mid-West Community Healthcare; Area 4, Cork Kerry Community Healthcare; Area 5, South East Community Healthcare; Area 6, Community Healthcare East; Area 7, Community Healthcare Dublin South, Kildare & West Wicklow; Area 8, Midlands Louth Meath Healthcare; Area 9, Dublin North City and County Healthcare

### Dietitians’ characteristics and experience of delivering weaning workshops

Data for delivery of weaning workshops were available for 30 participants, with most dietitians reporting not having delivered weaning workshops (n=19, 63.3%) (Table 2). Of the dieticians that had personally delivered weaning workshops previously, most were aged between 41-50 years (n=6, 54.6%), based in primary care (n=9, 81.8%) and in a senior dietitian’s role (n=9, 81.8%). Characteristics were mostly similar for dietitians who had not delivered workshops previously, with the majority aged between 31-40 years (n=8, 42.1%), based in primary care (n=14, 73.7%) and in a senior dietitian role (n=14, 73.7%). The findings of chi-squared tests are presented in Tables 2-4, with data for dieticians with experience of delivery presented separately to those without experience.

**Table 2.** Dietitians’ characteristics and delivery of weaning workshops.

|  | n | Delivered a WW (n=30) |  |
| --- | --- | --- | --- |
|  |  | Yes (n=11, 36.7%) | No (n=19, 63.3%) |
| **Age category** | 30 |  |  |
| 18-30 years |  | 1 (9.1) | 3 (15.8) |
| 31-40 years |  | 4 (36.4) | 8 (42.1) |
| 41-50 years |  | 6 (54.6) | 6 (31.6) |
| 51-60 years |  | 0 (0.0) | 1 (5.3) |
| >60 years |  | 0 (0.0) | 1 (5.3) |
| **Gender** | 30 |  |  |
| Female |  | 11 (100.0) | 18 (94.7) |
| Male |  | 0 (0.0) | 1 (5.3) |
| **CHO** | 30 |  |  |
| Area 1 |  | 2 (18.2) | 1 (5.3) |
| Area 2 |  | 0 (0.0) | 0 (0.0) |
| Area 3 |  | 0 (0.0) | 3 (15.8) |
| Area 4 |  | 2 (18.2) | 4 (21.1) |
| Area 5 |  | 0 (0.0) | 4 (21.1) |
| Area 6 |  | 2 (18.2) | 4 (21.1) |
| Area 7 |  | 2 (18.2) | 1 (5.3) |
| Area 8 |  | 0 (0.0) | 0 (0.0) |
| Area 9 |  | 3 (27.3) | 2 (10.5) |
| **Length of time in profession** | 30 |  |  |
| 1-5 years |  | 2 (18.2) | 9 (47.4) |
| 6-10 years |  | 2 (18.2) | 2 (10.5) |
| 11-15 years |  | 3 (27.3) | 4 (21.1) |
| 16-20 years |  | 4 (36.4) | 4 (21.1) |
| 21-25 years |  | 0 (0.0) | 0 (0.0) |
| **Discipline** | 30 |  |  |
| Primary care |  | 9 (81.8) | 14 (73.7) |
| Health promotion |  | 2 (18.2) | 2 (10.5) |
| Other |  | 0 (0.0) | 3 (15.8) |
| **Current position** | 30 |  |  |
| Dietitian manager |  | 1 (9.1) | 1 (5.3) |
| Senior dietitian |  | 9 (81.8) | 14 (73.7) |
| Staff-grade dietitian |  | 1 (9.1) | 4 (21.1) |
| **Parents looking for support** | 27 |  |  |
| Daily |  | 2 (18.2) | 0 (0.0) |
| Once a week |  | 4 (36.4) | 4 (25.0) |
| Once a month |  | 3 (27.3) | 6 (37.5) |
| Few times a year |  | 2 (18.2) | 2 (12.5) |
| Never |  | 0 (0.0) | 4 (25.0) |

WW, weaning workshop; CHO, Community Health Organisation

### Dietitians’ perceptions of workshop attendees

The majority of dietitians who have delivered weaning workshops (n=9, 81.8%) reported that parents who attend weaning workshops are mostly of medium socioeconomic status. Of those who had delivered weaning workshops, the majority indicated that only mothers (n=9, 81.8%) with their infants (n=11, 100.0%), aged 4-7 months (n=8, 72.9%) attended weaning workshops. Most dietitians who had not delivered a weaning workshop answered ‘I don’t know’ to most of the questions including ‘who attends weaning workshop’s (n=7, 53.9%), ‘parents’ socioeconomic status’ (n=6, 50.0%), ‘how many attended a weaning workshop class’ (n=5, 45.5%) and ‘how parents were referred’ to a weaning workshop (n=5, 45.5%). Finally, the majority of dietitians who had delivered weaning workshops reported attendees’ feedback as ‘excellent’ (n=7, 63.6%) (Table 3).

**Table 3.** Dietitians’ perceptions of those attending weaning workshops and delivery of weaning workshops.

| Variable | n | Delivered a WW (n=30) |  |
| --- | --- | --- | --- |
|  |  | Yes (n=11, 36.7%) | No (n=19, 63.3%) |
| ***How many attend WW (n=26)*** | 22 |  |  |
| ≤10 participants |  | 5 (45.5) | 4 (36.4) |
| >10 participants |  | 6 (54.6) | 2 (18.2) |
| Don't know |  | 0 (0.0) | 5 (45.5) |
| ***Who attends WW (n=28)*** | 24 |  |  |
| Both mother and father |  | 2 (18.2) | 0 (0.0) |
| Mother only |  | 9 (81.8) | 6 (46.2) |
| Don't know |  | 0 (0.0) | 7 (53.9) |
| ***Parents SES (n=26)*** | 23 |  |  |
| High |  | 0 (0.0) | 2 (16.7) |
| Medium |  | 9 (81.8) | 4 (33.3) |
| Low |  | 0 (0.0) | 0 (0.0) |
| Don't know |  | 2 (18.2) | 6 (50.0) |
| ***Are infant present at WW (n=35)*** | 30 |  |  |
| Yes |  | 11 (100.0) | 7 (36.8) |
| No |  | 0 (0.0) | 7 (36.8) |
| Don't know |  | 0 (0.0) | 5 (26.3) |
| ***Approximation of infant age (n=24)*** | 21 |  |  |
| 1-3 months |  | 0 (0.0) | 0 (0.0) |
| 4-7 months |  | 8 (72.9) | 5 (50.0) |
| 8+ months |  | 0 (0.0) | 0 (0.0) |
| Don't know |  | 3 (27.3) | 5 (50.0) |
| ***How are they referred to WW (n=25)*** | 22 |  |  |
| Self-referral |  | 2 (18.2) | 2 (18.2) |
| GP referral |  | 0 (0.0) | 0 (0.0) |
| Public health nurse |  | 5 (45.5) | 4 (36.4) |
| Community dietitian referral |  | 0 (0.0) | 0 (0.0) |
| Other |  | 4 (36.4) | 5 (45.5) |
| ***Perceptions of attendees' feedback (n=12)*** | 12 |  |  |
| Excellent |  | 7 (63.6) | 0 (0.0) |
| Very good |  | 3 (27.3) | 1 (100.0) |
| Good |  | 1 (9.1) | 0 (0.0) |
| Poor |  | 0 (0.0) | 0 (0.0) |
| Very poor |  | 0 (0.0) | 0 (0.0) |

WW, weaning workshops

### Weaning workshop format, content, development, and delivery of weaning workshops

Of the eleven dietitians who had previously delivered a workshop, 36.4% (n=4) reported that workshops were delivered by community dieticians in collaboration with public health nurses, while 45.5% did not know what other healthcare professionals were also involved in delivering weaning workshops (Table 4). Of dietitians with experience of delivering weaning workshops, 54.6% reported that workshop content included discussion on the timing of introduction of solid foods; responding to infant cues; stages of food textures, allergies, and baby-led weaning (n=6). According to the dietitians with experience, weaning workshops were evaluated using either informal participant feedback (n=3, 27.3%) or formal evaluation of opinions (n=3, 27.3%). For those who had delivered weaning workshops, the majority (n=4, 44.4%) mostly indicated that the time taken to travel to workshops was approximately 21-40 minutes, and to prepare for the workshops was 41-60 minutes. Most dietitians who had not delivered a weaning workshop did not know how weaning workshops were developed (n=16, 84.2%) or promoted (n=4, 36.6%), with some indicating that promotion was through other healthcare professionals (n=6, 54.6%). When asked about the cost of weaning workshops in terms of resources needed such as room hire or equipment, of those who had delivered weaning workshops, 36.4% reported no cost (n=4), with 18.2% reporting €101-€200 (n=2).

**Table 4.** Weaning workshop format, content, development, and delivery of weaning workshops.

| Variable | n | Delivered a WW (n=30) |  |
| --- | --- | --- | --- |
|  |  | Yes (n=11, 36.7%) | No (n=19, 63.3%) |
| ***Who delivers WW (n=25)*** | **21** |  |  |
| Community dietitian only |  | 1 (9.1) | 0 (0.0) |
| Public Health Nurse only |  | 1 (9.1) | 6 (60.0) |
| Community dietitian and public nurse |  | 4 (36.4) | 2 (20.0) |
| Don't know |  | 5 (45.5) | 2 (20.0) |
| ***Frequency of WW (n=40)*** | **30** |  |  |
| Once a month |  | 4 (36.4) | 4 (21.1) |
| Once a fortnight |  | 0 (0.0) | 0 (0.0) |
| Every 4 months |  | 2 (18.2) | 0 (0.0) |
| Twice a year |  | 1 (9.1) | 0 (0.0) |
| Never |  | 0 (0.0) | 8 (42.1) |
| Don't know |  | 4 (36.4) | 7 (36.8) |
| ***Topics discussed at WW (n=24)*** | **21** |  |  |
| Timing of introduction of solid foods |  | 0 (0.0) | 0 (0.0) |
| Responding to infant cues |  | 0 (0.0) | 0 (0.0) |
| Stages of food textures |  | 2 (18.2) | 3 (30.00) |
| Allergies |  | 0 (0.0) | 0 (0.0) |
| Baby-led weaning |  | 0 (0.0) | 1 (10.0) |
| All the above |  | 6 (54.6) | 0 (0.0) |
| Other |  | 3 (27.3) | 1 (10.0) |
| Don't know |  | 0 (0.0) | 5 (50.0) |
| ***Resources used at WW (n=23)*** | **21** |  |  |
| Videos |  | 0 (0.0) | 0 (0.0) |
| Power point slides |  | 0 (0.0) | 0 (0.0) |
| Cooking demo |  | 1 (9.1) | 1 (10.0) |
| Healthy Ireland 'Feeding your baby' booklet |  | 2 (18.2) | 2 (20.0) |
| 101 Square Meals Cookbook |  | 0 (0.0) | 0 (0.0) |
| Self-developed booklets/leaflets |  | 0 (0.0) | 0 (0.0) |
| Food display |  | 0 (0.0) | 0 (0.0) |
| All the above |  | 1 (9.1) | 0 (0.0) |
| Other |  | 7 (63.6) | 0 (0.0) |
| Don't know |  | 0 (0.0) | 7 (70.0) |
| ***How WW are developed (n=30)*** | **30** |  |  |
| Self-developed |  | 0 (0.0) | 0 (0.0) |
| By HSE primary care dietitian in your area |  | 3 (27.3) | 0 (0.0) |
| By another HCP |  | 1 (9.1) | 2 (10.5) |
| Adapted from another HSE area |  | 4 (36.4) | 1 (5.3) |
| Don't know |  | 3 (27.3) | 16 (84.2) |
| ***How WW are promoted (n=25)*** | **22** |  |  |
| Social media |  | 0 (0.0) | 0 (0.0) |
| Community centre |  | 1 (9.1) | 1 (9.1) |
| Suggested by HCPs |  | 6 (54.6) | 6 (54.6) |
| Other |  | 4 (36.6) | 0 (0.0) |
| Don't know |  | 0 (0.0) | 4 (36.6) |

**Table 4. Weaning workshop format, content, development, and delivery of weaning workshops** 569 **(continued)**
| Variable | n | Delivered a WW (n=30) |  |
| --- | --- | --- | --- |
|  |  | Yes (n=11, 36.7%) | No (n=19, 63.3%) |
| ***How WW are evaluated (n=19)*** | 19 |  |  |
| Informal participant feedback |  | 3 (27.3) | 0 (0.0) |
| Formal evaluation of opinions |  | 3 (27.3) | 0 (0.0) |
| Formal evaluation of weaning knowledge |  | 1 (9.1) | 0 (0.0) |
| Formal evaluation of weaning self-efficacy |  | 1 (9.1) | 0 (0.0) |
| Other |  | 3 (27.3) | 0 (0.0) |
| Don't know |  | 0 (0.0) | 8 (100.0) |
| ***Cost of WW(n=21)*** | 21 |  |  |
| No cost |  | 4 (36.4) | 2 (20.0) |
| €0-50 |  | 0 (0.0) | 0 (0.0) |
| €51-100 |  | 0 (0.0) | 0 (0.0) |
| €101-200 |  | 2 (18.2) | 0 (0.00) |
| €201-500 |  | 0 (0.0) | 0 (0.0) |
| Other |  | 2 (18.2) | 0 (0.0) |
| Don't know |  | 2 (18.2) | 8 (80.0) |
| ***Time to travel to WW (n=10)*** | 10 |  |  |
| 0-10 mins |  | 3 (3.3) | 1 (100.0) |
| 11-20 mins |  | 0 (0.0) | 0 (0.0) |
| 21-40 mins |  | 4 (44.4) | 0 (0.0) |
| 41-60 mins |  | 2 (22.2) | 0 (0.0) |
| >60mins |  | 0 (0.0) | 0 (0.0) |
| Don't know |  | 0 (0.0) | 0 (0.0) |
| ***Time to prepare for WW (n=12)*** | 12 |  |  |
| 0-10 mins |  | 2 (18.2) | 0 (0.0) |
| 11-20 mins |  | 2 (18.2) | 1 (100.0) |
| 21-40 mins |  | 2 (18.2) | 0 (0.0) |
| 41-60 mins |  | 4 (36.4) | 0 (0.0) |
| >60mins |  | 1 (9.1) | 0 (0.0) |
| Don't know |  | 0 (0.0) | 0 (0.0) |
| ***Time to close WW (n=11)*** | 11 |  |  |
| 0-10 mins |  | 3 (30.0) | 0 (0.0) |
| 11-20 mins |  | 5 (50.0) | 1 (100.0) |
| 21-40 mins |  | 2 (20.0) | 0 (0.0) |
| 41-60 mins |  | 0 (0.0) | 0 (0.0) |
| >60mins |  | 0 (0.0) | 0 (0.0) |
| Don't know |  | 0 (0.0) | 0 (0.0) |
| ***Other time needed for WW (n=4)*** | 4 |  |  |
| 0-10 mins |  | 0 (0.0) | 0 (0.0) |
| 11-20 mins |  | 2 (66.7) | 0 (0.0) |
| 21-40 mins |  | 0 (0.0) | 0 (0.0) |
| 41-60 mins |  | 0 (0.0) | 0 (0.0) |
| >60mins |  | 1 (33.3) | 0 (0.0) |
| Don't know |  | 0 (0.0) | 1 (100.0) |

WW, weaning workshops; HCP, health care professional: HSE, Health Service Executive

### Factors influencing the development and delivery of workshops

Participants identified resource constraints and a lack of staffing as primary reasons that workshops were not currently offered within their area. Participants also identified several factors they felt should be considered in the future development and delivery of weaning workshops. These include workshop characteristics such as appropriate content, timing, venue and mode of delivery, as well as organizational characteristics such as the availability of resources, multidisciplinary team (MDT) involvement, and attendee characteristics. One participant felt that weaning workshops were not needed within primary care. This data is further described in Table 5 which also provides the numbers of participants who identified each factor and sample quotes.

**Table 5.** Factors to consider in the development and delivery of workshops.

| Category | Factors<br>(n=number of respondents who identified this) | Example quotes |
| --- | --- | --- |
| Workshop characteristics | **Content:**<br>Practical and interactive (n=5)<br>Standardised/structured (n=4)<br>Evidence-based (n=3)<br>Responsive feeding/industry influence/stages of textures (n=3) | 'Any program should be interesting and interactive'<br>'Using existing materials to avoid duplication & save time, standardisation of material if possible'<br>'Nationally standardised weaning workshops available online and offline should be available for new parents and carers'<br>'Very important that there is more out there for parents/ families re weaning advice - especially evidence-based and safe advice!'<br>'We also need to ensure messages around responsive feeding are shared with parents'<br>'Nationally agreed referral pathway with a structured programme incorporated as part of the dietetic service to primary care'<br>'Most important component re weaning is to make sure textures are introduced in a timely fashion. We see many fussy eaters who took pureed foods for far too long' |
|  | **Timing:**<br>Appropriate to child age/stage (n=5) | 'To have them running a few times a year, to ensure you are able to see families at appropriate times'<br>'National weaning program - offered to all parents when their baby is between 3 and 6 months' |
|  | **Venue:**<br>Appropriate baby-friendly location (n=7) | 'Because Mums attend with children it is necessary to have the workshop in a room that caters for children' |
|  | **Mode of delivery:**<br>Accessible and free (n=4)<br>Use of peer leaders (n=1) | 'That they are accessible to all and free of charge'<br>'I do enjoy delivering the workshops myself, but appreciate that to scale up the trained [peer] tutor led approach would work best' |
| Resources | Staff time/availability (n=8)<br>Staff training/resources (n=3)<br>Administration support (n=2) | 'Dedicated committed staff so it won't flounder...a few key people driving it'<br>'Staffing such as releasing staff to deliver and continue to invest and develop the programme...allow staff to gain skills such as facilitation'<br>'Adequate funding of the project is essential to its continued success' |
| Multidisciplinary team (MDT) involvement | • MDT involvement to facilitate recruitment/referral (n=7)<br>• MDT involvement in development/delivery (n=5) | • *'It needs to be inclusive of all relevant disciplines not just Public Health Nursing driven and led'*<br>• *'Joint delivery between PHN and dietitian works well - has strengthened the relationship between nursing and dietetics'*<br>• *'Could be alternated between PHN, peer leader and Dietitian, would need PHN buy in'* |
| Participant characteristics | • Engaging participants from low SES (n=6)<br>• Participant attendance (n=2) | • *'How to engage those most in need of the information. Over the past 19 years it is middle-class well-informed parents that attend these workshops'*<br>• *'Access for lower socio-economic groups. They are under such social stress that they don't take up groups/workshops.'*<br>• *'The mums that attend is generally very interested in good nutrition and probably have read widely on the subject prior to attending. Unfortunately, it is difficult to get the mums who really need education on baby weaning to attend'* |

Overall, many participants regardless of their experience of delivery highlighted that workshops should be standardized and structured, and that content and associated materials should be practical and interactive, evidence-based and cover topics like responsive feeding, the potential influence of the infant feeding industry and stages of textures. For example, one participant wrote that there is a need for a *‘nationally agreed referral pathway with a structured programme incorporated as part of the dietetic service to primary care’*. Dietitians also identified that workshops should be appropriate to the child age/stage, e.g., to be run a few times a year to engage with parents and caregivers at appropriate times, and be held in child-friendly venues, at no cost to participants with one stating ‘*because Mums attend with children it is necessary to have the workshop in a room that caters for children’*. Regarding the resources needed to develop and implement these workshops, participants identified that availability of staff including administrative support was crucial. They also stated that MDT involvement was crucial both in terms of recruiting or referring attendees to the workshop, and in the development and delivery of workshops, as one participant remarked, *‘it needs to be inclusive of all relevant disciplines’’*. Finally, in relation to attendee characteristics, dietitians stated the importance of running workshops for parents and/or caregivers with greatest need, e.g. with a particular focus on engaging families from lower socio-economic status backgrounds, *‘The mums that attend is [sic] generally very interested in good nutrition and probably have read widely on the subject prior to attending. Unfortunately, it is difficult to get the mums who really need education on baby weaning to attend’*.

## Discussion

This study provides an overview of current practice regarding the delivery of weaning workshops within the Irish primary healthcare service. The study also provides insight into the experiences and opinions of primary care community dietitians regarding the development and delivery of weaning workshops in terms of optimal content and delivery options. Overall, only 11 dietitians reported experience of personally delivering weaning workshops. Sixteen dietitians reported that workshops were run in their area with variable frequency, with ten reporting that workshops were never run in their area. Participants reported that mostly only mothers of medium socioeconomic status attended weaning workshops when infants were aged between 4-7 months, and that feedback from attendees was mostly positive. Dietitians identified the key factors needing to be considered in the future development and delivery of weaning workshops as workshop characteristics such as content, timing and venue, organisational characteristics such as the availability of resources and multidisciplinary involvement, and attendee characteristics such as socioeconomic status.

This study highlights substantial variability in current practice regarding weaning workshops in Irish primary care, with workshops being run in some areas by some dietitians but not in others, as well as a lack of standardisation regarding the provider, content, and frequency of workshops where workshops are being delivered. We also identified a distinct lack of clarity and knowledge amongst dietitians regarding what current practice is regarding weaning workshops, with many reporting ‘don’t know’ responses. However, participants agreed on several key factors to be considered for the future development and implementation of weaning workshops. Like other studies in this area, the timing of weaning workshops and ensuring appropriateness of the workshop to the child age/stage was a crucial factor identified by dietitians in this study. Previous research has indicated that mothers are more responsive to personalised approaches within infant feeding interventions (37, 38) and that tailoring workshops to target different stages of infant development is important to optimise their effectiveness (29). Previous intervention studies have indicated that interventions initiated antenatally (39) or up to 10 weeks after birth (40, 41) can be effective in promoting appropriate infant feeding (29). However, this study found that workshops currently offered involve infants between 4-7 months of age, with no dietitian reporting workshops delivered to parents with infants aged 0-3 months. Given the fact that infants are often introduced to solids before four months, and sometimes as early as 12 weeks as outlined earlier (3-7), this further highlights the importance of carefully considering the timing of such programmes. Weekly or fortnightly workshops with continuous advice provided for all stages of development might help to improve infant feeding practices and nutritional habits.

This study also highlights the importance of targeting weaning workshops and infant feeding programmes towards the areas of most need. A common theme identified by participants was the need to engage parents and caregivers from lower socioeconomic status backgrounds; however, dietitians also reported that workshops are currently predominantly attended by mothers of medium socioeconomic status. Existing guidance emphasises the importance of different intervention approaches which can focus on deprived groups (29) to target the recurring relationship that exists between socioeconomic status and inappropriate infant feeding practices (35, 42, 43). Previous studies have identified maternal education and socioeconomic status as factors which predict poor infant feeing behaviours (44, 45). Given that participants identified the availability of resources (e.g. staff time, cost) as a key factor influencing the development and delivery of weaning workshops, in resource constrained settings it is likely to be particularly important to prioritise the provision of workshops for parents and caregivers from more deprived backgrounds. A previous evaluation of infant-feeding workshops amongst women living in areas of high deprivation found that a relatively short education intervention can enhance women’s knowledge and understanding of weaning (29). While demographic factors are unmodifiable, there is scope to focus on and influence modifiable factors such as women’s beliefs about infant feeding (35, 46). Therefore, the future development and implementation of weaning workshops should consider important aspects such as accessibility and cost, providing workshops in appropriate locations at different times, free of charge that do not incur additional expenses on attendees (e.g. childcare, parking), with more concerted efforts to specifically engage with and target families of lower socioeconomic status.

Dietitians in this study perceived mothers’ feedback on these weaning workshops as ‘excellent’. Similarly, a study which evaluated infant-feeding workshops for women from deprived areas, found the workshops were generally rated positively while also being seen as an accessible source of education (29). Furthermore, several other studies found that after attending educational and practical workshops on infant feeding and complementary feeding women appeared to have improved knowledge, understanding and confidence with many following the current infant feeding recommendations (37, 38, 41). While data such as this is encouraging, future research should formally evaluate attendees’ experiences, levels of knowledge and infant feeding behaviours prior to and following attendance at weaning workshops to ensure that content is clear and comprehensive.

## Limitations

A main limitation of this survey is that information provided on parent/primary caregiver attendees is from dietitian self-report and as we did not collect data on time since delivering workshops, it may also be influenced by participant recall. We did not have the resources to also collect data from parent/caregivers, therefore future research should explore weaning workshops with attendees to obtain a more comprehensive understanding of the phenomenon in question. In items that allowed for multiple responses (‘tick all that apply’), variables were re-categorised for the analysis, easy interpretation and presentation of results (47). However, by doing this, some information is lost, so caution must be used when interpreting the results. ‘Tick all that apply’ answer format in questionnaires do not require respondents to differentiate between the answer responses, therefore a series of forced choice Yes/No questions should be used as recommended by Rasinski, Mingay, and Bradburn (1994) in future research (48). In addition, only 11 participants reported first-hand experience of delivering weaning workshops and as such, some responses may have been more directly informed by tacit experience of delivering weaning workshops than others. However, our chi-squared tests showed no observed patterns of difference between the overall responses of those with and without experience, and we felt it important to include the views of all participants, as some may have been involved in commissioning or organising rather than delivery or may have drawn on other relevant experience. We also felt it was important to include the views of all dietitians who might potentially be involved in the delivery of future workshops to gives a more inclusive overview of potential factors and barriers/facilitators to consider.

## Conclusions

This study provides a snapshot of current delivery of community-based weaning workshops within the Irish health service. Using a cross-sectional survey, community-based dietitians perceived participants’ feedback on weaning workshops as excellent, while highlighting issues around optimal timing and delivery of workshops. Furthermore, the findings highlight that future research needs to focus on how to engage families of lower socioeconomic status and the impact of these workshops on behaviour change, taking into consideration key factors such as accessibility and cost. Although based within an Irish setting, the need to support complementary feeding and subsequent rationale for establishing weaning workshops are extremely pertinent internationally. Given the dearth of evidence in this area, the insights provided by participants of this study regarding the development and delivery of weaning workshops, in terms of optimal content and delivery options, will make a valuable contribution to this field.

## Supporting information

Appendix 1 STROBE Checklist

Appendix 2 Questionnaire

Appendix 3 Missing data info

## Data Availability

All data produced in the present study are available upon reasonable request to the authors

## Acknowledgements

The authors would like to sincerely thank the participants who took the time to complete this survey. This research received no specific grant from any funding agency, commercial or not-for-profit sectors and was conducted as part of a MA in Public Health in University College Cork undertaken by Caroline Shea and supervised by Dr Elaine Toomey and Dr Caragh Flannery.

