## Appendix 2 Questionnaire for "Investigating group-based classes (‘weaning workshops’) to support complementary infant feeding in Irish primary care settings: A cross-sectional survey"

**Exploring current practice regarding weaning workshops in Ireland**

I am a student at the University College Cork pursuing a Masters of Public Health. My thesis aims to explore current practice in Ireland regarding weaning workshops. For the purposes of this survey, the term ‘weaning workshop’ will be used to describe any community-based, group-based classes that aim to address complementary feeding and the introduction of solid foods for parents and primary caregivers of infants and children.

A brief report on HSE weaning workshops previously identified a substantial amount of variability in terms of what weaning workshops currently exist. This survey aims to build on this previous report to identify past and current practice regarding weaning workshops delivered within the HSE, as well as briefly exploring the experiences and opinions of HSE community dietitians involved in developing and delivering weaning workshops. Results will provide an overview of past and current services and may be used to inform the development and standardization of future weaning workshops.

This survey should take approximately 10-15 minutes to complete. There are no right or wrong answers. Please answer these questions to the best of your ability.

This survey is completely confidential, anonymous, and voluntary.

**Please provide brief background information**

1. Please indicate your age below:

a. 18-20

b. 21-30

c. 31-40

d. 40-49

e. 50-59

f. 60 or older

2. To which gender identity do you most identify?

a. Male

b. Female

c. Transgender female

d. Transgender male

e. Prefer not to say

f. Other (please detail): ______________________________________________________________________

3. In which Community Health Organisation (CHO) are you based? Please also provide exact location e.g. Area 4: Glanmire/Macroom)

a. Area 1: ____________________________

b. Area 2: ____________________________

c. Area 3: ____________________________

d. Area 4: ____________________________

e. Area 5: ____________________________

f. Area 6: ____________________________

g. Area 7: ____________________________

h. Area 8: ____________________________

i. Area 9: ____________________________

4. In what year did you qualify as a dietitian?

_____________________________

5. How long have you been working in the community?

a. 1-5

b. 6-10

c. 11-15

d. 16-20

e. 21-25

f. 26-30

g. 31-35

h. 36-40

i. 40+

6. Is your role primary care or health promotion?

a. Primary care

b. Health promotion

7. Please indicate what best describes your current position:

a. Dietitian Manager

b. Senior Dietitian

c. Staff-grade Dietitian

d. Other (please detail): ______________________________________________________________________

**Please tell us about weaning workshops in your area**

8. How frequently are HSE-run weaning workshops held in your area?

a. Once a week

b. Once a fortnight

c. Once a month

d. Once every four months

e. Twice a year

f. Never

g. I don’t know

h. Other (please explain): ______________________________________________________________________

9. If HSE-run workshops are not being delivered in your area, please briefly explain why:

______________________________________________________________________________________

10. Are you aware of any non-HSE run weaning workshops in your area:

Yes

No

I don’t know

11. If yes, please briefly describe the non-HSE run workshops in your area (e.g. private dietician, peer-led etc.):

_______________________________________________________________________________

10. If applicable, please indicate the amount and duration of HSE-run weaning workshop	sessions in your area (e.g. 5 x 60- minute sessions)

_________________________________________________________

Not applicable

11. If applicable, please indicate who delivers HSE-run workshops?

a. Community dietitian only

b. Public Health Nurse only

c. Community dietitian and Public Health Nurse

d. Community dietitian and other (please explain):	________________________________________

e. Other (please explain): ____________________________________________________________

f. I don’t know

12. On average, how many participants attend these weaning workshops?

a. 0-5

b. 6-10

c. 11-15

d. 16-20

e. 21-25

f. 26-30

g. 30+

h.Other (please explain): __________________________________________________________________

i. I don’t know

13. On average, who attends these workshops? *Tick all that apply*

a. Mother

b. Father

c. Grandmother

d. Grandfather

e. Other (please explain): __________________________________________________________________

f. I don’t know

14. On average, what socioeconomic status do participants belong to?

a. Higher

b. Middle

c. Lower

d. I don’t know

15. Are infants/toddlers present for these workshops?

a. Yes

b. No

c. I don’t know

16. On average, what age are infants/toddlers when parents/caregivers first attend workshops?

a. 0-4 weeks

b. 5-8 weeks

c. 9-12 weeks

d. 13-16 weeks

e. 17-20 weeks

f. 21-24 weeks

g. 25-28 weeks

h. 29-32 weeks

i. 33 weeks+

j. Other (please explain): ______________________________________________________________________

k. I don’t know

17.	How are attendees referred to the workshop?

a. Self-referral

b. GP referral

c. Public Health Nurse referral

d. Community dietitian referral

e. Other (please explain):_________________________________________________________________

f. I don’t know

18. How are HSE-run weaning workshops promoted or advertised? *tick all that apply*

a. Social media
b. Community centres
c. Suggested by Healthcare Professional (HCP)
d. Other (please explain): ___________________________________________________________
e. I don’t know

19. What topics do the HSE-run weaning workshops in your area address? *Tick all that apply*

a. Timing of introduction of solid foods

b. Responding to infant cues

c. Stages of food textures

d. Allergies

e. Baby-led weaning

f. Other (please explain): __________________________________________________________________

g. Not applicable

h. I don’t know

20. What resources and materials are used to deliver HSE-run workshops in your area? *Tick all that apply.*

a. Videos
b. PowerPoint slides
c. Cookery demonstrations
d. Healthy Ireland ‘Feeding your baby’ booklet
e. 101 Square Meals Cookbook
f. Self-developed booklets/leaflets
g. Food display
h. Other (please explain): _____________________________________________________________
i. Not applicable
j. I don’t know

21. What resources and materials are provided to parents during HSE-run workshops?

a. Healthy Ireland ‘Feeding your baby’ booklet
b. 101 Square Meals Cookbook
c. Self-developed booklets/leaflets
d. Information on online resources and supports (please explain): ________________________________________________________________________________________
e. Information on local resources and supports (please explain): ________________________________________________________________________________________
f. Other (please explain): _____________________________________________________________
g. Not applicable
h. I don’t know

22. How was the workshop developed?

a. Self-developed

b. Developed by HSE primary care dietitian in your area

c. Developed by another HCP (please explain): __________________________________________

d. Adapted from another HSE area	e. Adapted from an existing international clinical	trial (e.g. INFANT study) (please explain):_____________________________________________________________

e. Other (please explain): _____________________________________________________________

f. Not applicable

g. I don’t know

23. How are the workshops evaluated?

a. Informal participant feedback

b. Formal evaluation of participant opinions

c. Formal evaluation of participant weaning knowledge

d. Formal evaluation of participant self-efficacy in relation to weaning

e. Other (please explain): _____________________________________________________________

f. Not applicable

g. I don’t know

24. On average, what is participant feedback towards workshops?

a. Excellent

b. Very Good

c. Good

d. Poor

e. Very poor

25. What is the estimated total cost of delivering HSE-run weaning workshops in your area in terms of resources needed (e.g. room hire, equipment etc.)?

a. No cost

b. 0-50e

c. 51-100e

d. 101-200e

e. 201-500e

f. Other (please explain): _____________________________________________________________

g. Not applicable

h. I don’t know

26. Please estimate how many minutes in total the workshop requires of staff time (excluding delivery)

a. Travelling to and from workshop: _________________________

b. Preparing for the workshop: _______________________________

c. Closing and clearing up afterwards:_______________________

d. Other time taken (please explain):________________________

_________________________________________________________________________________________________

**Please tell us about your experience and opinions of delivering weaning workshops**

27. Have you delivered HSE-run weaning workshops previously?

a. Yes

b. No

28. If yes, how many HSE-run weaning workshops have you held in the last 12 months?

29. How frequently do you encounter parents/caregivers of young children looking for weaning information/workshops for support?

a. Daily

b. Once a week

c. Once a month

d. A few times a year

e. Less than a few times a year

f. Never

g. I don’t know

30. What factors do you believe influence attendance at workshops by parents/caregivers?

a. Location of workshops

b. Cost

c. Time to attend

d. Relationships with peers (e.g. family and friends)

e. Relationships with HCPs referring

f. Relationships with HCP delivering workshop

g. Content of workshops

h. Socio-economic status

i. Advertising and promotion of workshops

j. Other (please explain): ________________________________________________________________

k. I don’t know

31. What factors do you believe influence the development and delivery of weaning workshops in your area?

a. Cost

b. Location of workshops

c. Time to prepare

d. Time to deliver

e. Relationships with HCPs referring

f. Other (please explain): ________________________________________________________________

g. I don’t know

32. What do you feel are the most important factors to consider in the development and delivery of weaning workshops in primary care?

___________________________________________________________________________________________________________________________________________________________________________________________________________________________________________________________________________________________________________________________

33. What changes, if any, do you think should be made to existing service delivery in regards to weaning workshops?

___________________________________________________________________________________________________________________________________________________________________________________________________________________________________________________________________________________________________________________________

34. Any other comments?

_________________________________________________________________________________________________________

__________________________________________________________________________________________________________________________________________________________________________________________________________________

*Thank you for your participation in this survey!*
