## Appendix 3 Missing data info for "Investigating group-based classes (‘weaning workshops’) to support complementary infant feeding in Irish primary care settings: A cross-sectional survey"

**Appendix 3 - Missing data for each variable**

| **Variable** | **Missing** | **Total** | **Missing Percent** |
| --- | --- | --- | --- |
| Age category | 4 | 47 | 8.5 |
| Gender | 5 | 47 | 10.6 |
| CHO | 5 | 47 | 10.6 |
| Length of time in profession | 7 | 47 | 14.9 |
| Discipline | 6 | 47 | 12.8 |
| Current position | 5 | 47 | 10.6 |
| Delivery WW | 17 | 47 | 36.2 |
| Parents looking for support | 20 | 47 | 42.6 |
| How many attend WW | 21 | 47 | 44.7 |
| Who attends WW | 19 | 47 | 40.4 |
| Parents SES | 21 | 47 | 44.7 |
| Infants present at WW | 12 | 47 | 25.5 |
| Infants age | 23 | 47 | 48.9 |
| Referred to WW | 22 | 47 | 46.8 |
| Attenders feedback | 35 | 47 | 74.5 |
| Who delivers WW | 22 | 47 | 46.8 |
| Frequency of WW | 7 | 47 | 14.9 |
| Topics discussed at WW | 23 | 47 | 48.9 |
| Resources used at WW | 24 | 47 | 51.1 |
| How WW are developed | 17 | 47 | 36.2 |
| How WW are promoted | 22 | 47 | 46.8 |
| How WW are evaluated | 28 | 47 | 59.6 |
| Cost of WW | 26 | 47 | 55.3 |
| Time to travel | 37 | 47 | 78.7 |
| Time to prepare | 35 | 47 | 74.5 |
| Time to close | 36 | 47 | 76.6 |
| Other time needed | 43 | 47 | 91.5 |

WW = weaning workshop
